# Impact of community treatment with ivermectin for the control of scabies on the prevalence of antibodies to *Strongyloides stercoralis* in children

**DOI:** 10.1101/2019.12.18.19015248

**Authors:** Michael Marks, Sarah Gwyn, Hilary Toloka, Christian Kositz, James Asugeni, Rowena Asugeni, Jason Diau, John M Kaldor, Lucia Romani, Michelle Redman-MacLaren, David MacLaren, Anthony W Solomon, David CW Mabey, Andrew C Steer, Diana Martin

**Affiliations:** Clinical Research Department, Faculty of Infectious and Tropical Diseases, London School of Hygiene & Tropical Medicine, Keppel Street, London, WC1E 7HT, United Kingdom; Hospital for Tropical Diseases, University College London Hospitals NHS Trust, London, WC1E 6JB, United Kingdom; Division of Parasitic Diseases and Malaria, Centers for Disease Control and Prevention, Atlanta, United States of America; Atoifi Adventist Hospital, Atoifi, Malaita Province, Solomon Islands; Kirby Institute, University of New South Wales, Sydney, Australia; College of Medicine and Dentistry, James Cook University, Cairns, Queensland, Australia; Department of Paediatrics, University of Melbourne, Melbourne, Victoria, Australia; Department of General Medicine, Royal Children’s Hospital, Melbourne, Victoria, Australia; Tropical Diseases Research Group, Murdoch Children’s Research Institute, Melbourne, Victoria, Australia

**Author notes:** **Author for Correspondence:** Michael Marks, Clinical Research Department, Faculty of Infectious and Tropical Diseases, London School of Hygiene & Tropical Medicine, Keppel Street, London, WC1E 7HT, United Kingdom.

**Keywords:** Scabies, Neglected tropical diseases, Ivermectin, Strongyloides

## Abstract

Prevalence of antibodies to *Strongyloides stercoralis* was measured in 0–12-year-olds using a bead-based immunoassay before and after ivermectin mass drug administration (MDA) for scabies in the Solomon Islands. Seroprevalence was 9.3% before and 5.1% after MDA (p = 0.019), demonstrating collateral benefits of scabies MDA in this setting.

## Introduction

*Strongyloides stercoralis*, is unique among soil-transmitted helminths (STHs) in its ability to complete its life cycle within the human host. *S. stercoralis* infection is most commonly asymptomatic but may be associated with eosinophilia, fatigue, diarrhoea and occasionally larva currens [1]. People with compromised immune systems are at risk of potentially fatal hyperinfection syndrome [1]. Albendazole or mebendazole are effective against other major STH species and have been incorporated into mass drug administration (MDA) programs for public health purposes. However, these drugs have limited efficacy for strongyloidiasis, for which ivermectin is the first-line agent.

MDA with ivermectin is used widely to control a number of neglected tropical diseases (NTDs), including lymphatic filariasis, onchocerciasis and, most recently, scabies [2,3], but has so far not been adopted for *S. stercoralis* control. Given the drug’s broad anti-parasitic effect, MDA with ivermectin may have as a collateral benefit the population-level control of *S. stercoralis*. Evaluation of *S. stercoralis* control has been facilitated by serological assays that detect antibodies against the NIE antigen, which is present in infective L3 larvae [4] These antibodies likely indicate current or recent infection [5] and have high sensitivity and specificity compared to stool examination[4]. The NIE antigen has been adapted for use in the Luminex platform, allowing large scale screening of populations using dried blood spots [6].

In the context of a community-randomized trial evaluating the addition of azithromycin to ivermectin-based MDA for scabies and impetigo in the Solomon Islands, we measured the prevalence of antibody responses to the *S. stercoralis* NIE antigen before and after MDA.

## Methods

The trial of MDA for scabies and impetigo has been described elsewhere [7]. Briefly, selected communities in Malaita province in the Solomon Islands were randomised to MDA with open label ivermectin or ivermectin plus azithromycin. All residents of these communities were eligible to participate. In both trial arms, all participants were examined for scabies and offered a single oral dose of ivermectin (200 μg/kg of body weight). People with a contraindication to ivermectin (pregnancy, breast-feeding, or weight <15 kg) were offered topical permethrin instead. Those in whom a clinical diagnosis of scabies was made at baseline were given a second dose of ivermectin 7–14 days later. Written informed consent was obtained from adults and from a parent or guardian of each child aged under 18 years. Assent was also obtained from children who were able to provide it. The study was approved by the London School of Hygiene & Tropical Medicine Ethics Committee, the Solomon Islands National Health Ethics Committee and the Atoifi Adventist Hospital Ethics Committee. The main trial was prospectively registered on clinicaltrials.gov (NCT02775617). CDC staff did not interact with study participants or have access to identifying information.

For the sub-study reported here, we collected dried-blood spots (DBS) from all children aged less than 13 years at the baseline and 12-month surveys. We used a fluorescent bead-based assay to test for antibodies against the recombinant NIE antigen [6]. Briefly, serum was incubated with microspheres conjugated to NIE, beads were washed to remove unbound immunoglobulin (Ig), and then bound anti-NIE antibody was detected using biotinylated anti-human IgG+IgG4 antibody followed by streptavidin-phycoerythrin. Plates were run on a Luminex-200 (Austin, TX USA) and results reported as median fluorescence intensity with background subtracted. We used a receiver operating characteristic curve analysis to determine cut-offs for seropositivity.

We conducted a before and after analysis to determine the effect of the MDA on prevalence of antibodies to *S. stercoralis*. Azithromycin has no known activity against *S. stercoralis* so we combined the two trial arms into a single group for analysis. We calculated the seroprevalence of *S. stercoralis* at baseline and twelve months, and the absolute and relative reduction at twelve months, along with associated confidence intervals and statistical tests of significance. Statistical analysis was conducted in R 3.4.2 (Vienna, Austria).

## Results

In total, 1,291 people, including 553 0–12-year-olds were recruited and offered treatment for scabies at the time of the baseline survey. We collected DBS from 539 of these children, representing more than 97% of children enrolled in the trial. At the 12-month follow-up, 1,085 individuals (including 479 0–12-year-olds) were seen, and we collected DBS from 448 children (94%). At baseline, 9.3% of the children were seropositive for antibodies to NIE, with a range across the six study communities of 2.2% to 14.3% (Table 1). At the 12-month follow-up, the overall prevalence had declined to 5.1% with a range across the communities of 2.1% to 6.7%. The absolute difference in prevalence between baseline and 1 year was 4.2% (95% CI 0.7–7.5%) and the relative reduction was 45% (p = 0.019) The seroprevalence of antibodies to NIE was lower at the follow-up visit than at baseline in all communities (Table). Neither the baseline seroprevalence nor the magnitude of change following MDA was higher in communities with a higher baseline prevalence of scabies where more individuals would have received two doses of ivermectin at baseline (Table).

**TABLE 1:**
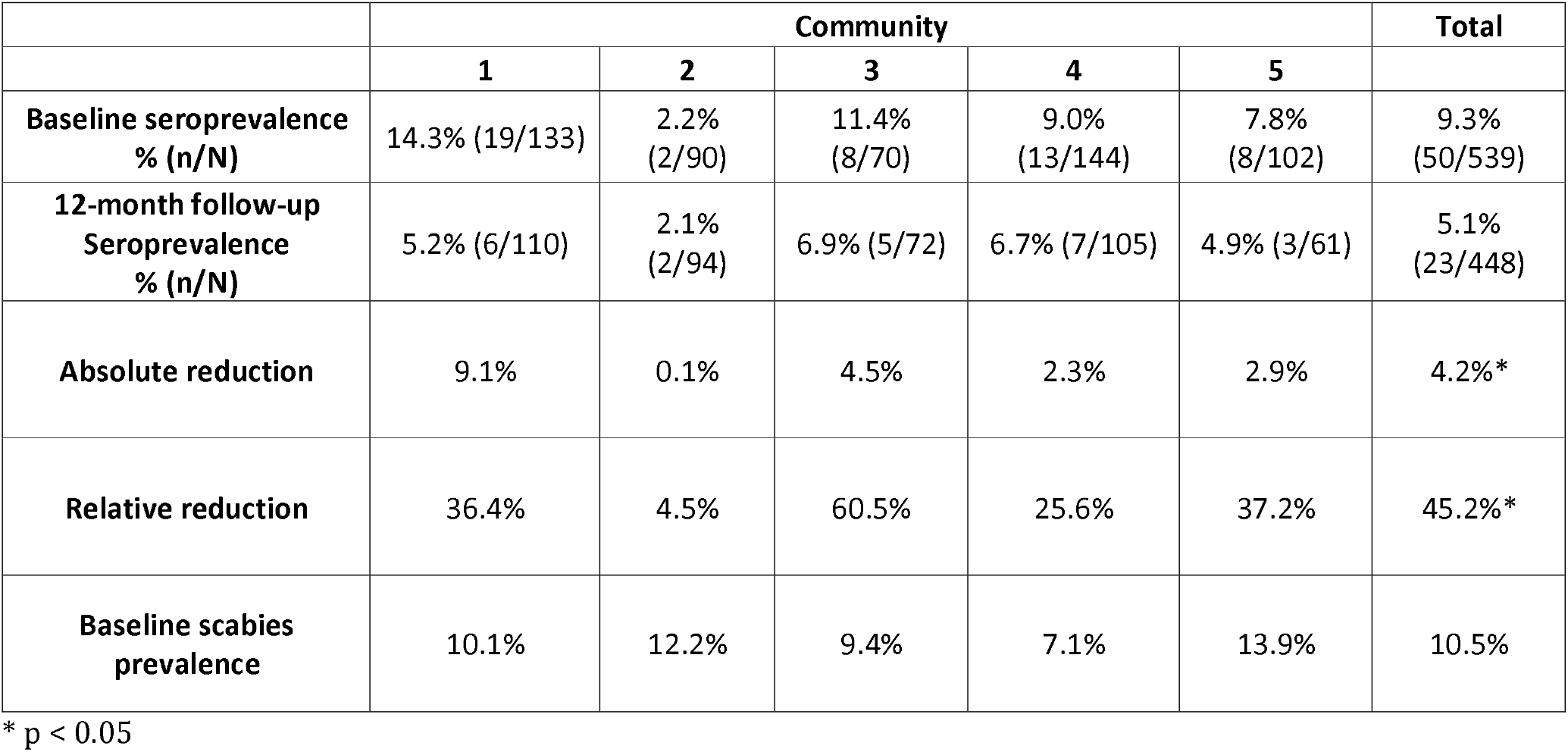
*Strongyloides stercoralis* seroprevalence in children aged 0–12 before and after ivermectin mass drug administration

## Discussion

Our study adds to the limited available data on the impact of ivermectin MDA on *S. stercoralis* prevalence. In the Northern Territory of Australia, a single round of MDA with ivermectin reduced *S. stercoralis* seroprevalence from 21% at baseline to 5% at 6 months, and a second MDA at month 12 further reduced the seroprevalence to 2% at month 18 [8]. In Ecuador, the prevalence of *S. stercoralis* fell from 6.8% to zero following multiple rounds of MDA conducted for the purpose of onchocerciasis elimination[9]. We showed that in 6 communities in Malaita, Solomon Islands, there was a decrease in seroprevalence of anti-NIE antibodies one year after a single ivermectin MDA conducted for the purpose of scabies control.

In the absence of a gold standard test for diagnosis of *S. stercoralis*, we used serological markers that correlate well with more direct measures of infection. Had we combined serology with other tests such as Kato-Katz or PCR on stool specimens, we may have detected more cases. We were, however, consistent in the measurement of exposure at both time points so are confident in the relative change observed. As complete seroreversion of anti-NIE responses after treatment may take more than one year[5], and it is not clear that all anti-NIE positive individuals will serorevert [6], it is possible that we have under-estimated the community-level effect of the intervention. Our study was not powered to detect changes in seroprevalence in each individual community, and although the changes were not statistically significant, there were declines in seroprevalence in every community, consistent with our overall study finding.

Data on the prevalence and distribution of STHs in the Solomon Islands are limited. Two previous studies showed that hookworm, whipworm and roundworm are common [10]. The baseline seroprevalence of *S. stercoralis* in 0– 12-year-olds in the current study was 9.3%, which is broadly similar to prevalence of other STH species reported in the Solomon Islands [10]. Current national deworming guidelines for the Solomon Islands are based on MDA with albendazole, which is likely to have little or no impact on *S. stercoralis*. Integrated strategies combining albendazole with ivermectin may therefore be beneficial for control of *S. stercoralis* as well as providing enhanced effectiveness against several STH species and allowing simultaneous control of scabies, which is highly endemic in the region [3,7]. The current requirement for two doses a of ivermectin, a week apart, for scabies MDA may represent a logistical barrier to integration as single-dose treatment is effective against *S. stercoralis* [11]. Newer anti-parasitic drugs such as moxidectin show promise as single-dose treatment for scabies, strongyloidiasis and other STH [12,13].

We have demonstrated that *S. stercoralis* is endemic in the Solomon Islands and added to the limited data demonstrating that ivermectin MDA, here conducted for the purpose of scabies control, can also reduce *S. stercoralis* seroprevalence. These data highlight ancillary benefits that can be conferred by NTD control programmes and suggest that STH programmes in this region may benefit from the addition of ivermectin alongside albendazole

## Data Availability

Data available in supplementary material

## Disclaimer

The authors alone are responsible for the views expressed in this article and they do not necessarily represent the views, decisions or policies of the institutions with which they are affiliated.

## Contributors

MM wrote the first draft of the paper. CB, KA, EP conducted laboratory work. MM, HT, CK, JA, RA conducted fieldwork. MM, CB, KA, analysed the data. MM, JD, JKM, LR, MRM, DM, AWS, DCWM, AS designed and supervised the study. All authors revised the manuscript.

## Supplementary Files

S1 Data – Study Dataset

S2 Checklist – Strobe Checklist

